# Diagnostic performance and prediction of clinical progression of plasma phospho-tau181 in the Alzheimer’s Disease Neuroimaging Initiative

**DOI:** 10.1101/2020.07.15.20154237

**Authors:** Thomas K. Karikari, Andréa L. Benedet, Nicholas J. Ashton, Juan Lantero Rodriguez, Anniina Snellman, Marc Suárez-Calvet, Paramita Saha Chaudhuri, Firoza Lussier, Hlin Kvartsberg, Alexis Moscoso Rial, Tharick A. Pascoal, Ulf Andreasson, Michael Schöll, Pedro Rosa-Neto, Kaj Blennow, Henrik Zetterberg, for the Alzheimer’s Disease Neuroimaging Initiative

## Abstract

Whilst cerebrospinal fluid (CSF) and positron emission tomography (PET) biomarkers for amyloid-β (Aβ) and tau pathologies are accurate for the diagnosis of Alzheimer’s disease (AD), their broad implementation in clinical and trial settings are restricted by high cost and limited accessibility. Plasma phosphorylated-tau181 (p-tau181) is a promising blood-based biomarker that is specific for AD, correlates with cerebral Aβ and tau pathology, and predicts future cognitive decline. In this study, we report the performance of p-tau181 in >1,000 individuals from the Alzheimer’s Disease Neuroimaging Initiative (ADNI), including cognitively unimpaired (CU), mild cognitive impairment (MCI) and AD dementia patients characterized by Aβ PET. We confirmed that plasma p-tau181 is increased at the preclinical stage of Alzheimer and further increases in MCI and AD dementia. Individuals clinically classified as AD dementia but having negative Aβ PET scans did not show increased plasma p-tau181. Despite being a multicenter study, plasma p-tau181 demonstrated high diagnostic accuracy to identify AD dementia (AUC=85.3%; 95% CI, 81.4%-89.2%), as well as to distinguish between Aβ- and Aβ+ individuals along the Alzheimer’s *continuum* (AUC=76.9%; 95% CI, 74.0%-79.8%). Higher baseline concentrations of plasma p-tau181 accurately predicted future dementia and performed comparably to the baseline prediction of CSF p-tau181. Longitudinal measurements of plasma p-tau181 revealed low intra-individual variability, which could be of potential benefit in disease-modifying trials seeking a measurable response to a therapeutic target. This study adds significant weight to the growing body of evidence in the use of plasma p-tau181 as a non-invasive diagnostic and prognostic tool for AD, regardless of clinical stage, which would be of great benefit in clinical practice and a large cost-saving in clinical trial recruitment.

## Introduction

Alzheimer’s disease (AD) is the most common form of neurodegenerative dementia and is defined by the accumulation of amyloid beta (Aβ) and tau aggregates in the brain [1]. These pathological changes occur several years before the manifestation of clinical symptoms [2] and are initiated by the build of extracellular amyloid beta (Aβ) plaques, followed by the accumulation of aggregated phosphorylated tau (p-tau) in the form of paired helical filaments in dystrophic neurites surrounding the plaques and in intraneuronal tangles [3]. At the symptomatic phase of the disease, Aβ is often widespread while the neuroanatomic distribution of tau tangles is more associated with the cognitive domains affected in patients with AD dementia [4].

AD dementia is most typically diagnosed based on clinical criteria; however, Aβ pathology can now be accurately measured using Aβ positron emission tomography (PET) or cerebrospinal fluid (CSF) Aβ42/Aβ40 ratio [5]. Tau pathology can also be detected by tau PET or CSF concentration of tau phosphorylated at threonine-181 (p-tau181), which are both highly specific for AD [6]. The downstream effects of Aβ and tau pathologies on neurodegeneration can be measured by magnetic resonance imaging (MRI), as well as CSF total-tau (t-tau) and neurofilament light (NfL). Neuroimaging and CSF biomarkers of AD are highly accurate for diagnosis and progression monitoring [7–9], however, these biomarkers have significant drawbacks for routine use in clinical settings. They are invasive, time-consuming and expensive, furthermore they are difficult to access, particularly in a primary care setting [10]. Blood biomarkers may hold promise to address the challenges of these current methods.

Methods for the detection of Aβ peptides in blood are available, the results of which are related to cerebral Aβ pathology [11–13]. However, extracerebral expression of Aβ peptides challenges their use in clinical laboratory practice. Blood biomarkers for tau have been lacking. While ultrasensitive plasma t-tau assays can detect neuronal injury in acute brain disorders, *e.g*., stroke and traumatic brain injury, they work relatively poorly in AD settings, and the correlation with CSF t-tau is weak [14]. Assays for the quantification of p-tau181 in blood have been recently developed and the results showed increased concentrations in AD dementia [15–18]. In cross-sectional single-center studies, blood p-tau181 was increased across the Alzheimer’s clinical *continuum* – from Alzheimer to AD dementia – and enabled the differential diagnosis of AD as compared to non-AD neurodegenerative disorders [15, 17, 19–21]. Furthermore, plasma p-tau181 correlates strongly with CSF p-tau181 and PET measures of Aβ and tau pathologies [15, 17, 19, 20]. However, routine clinical applications of plasma p-tau181 biomarkers would require a demonstration of robust validation in large multicenter studies. Furthermore, whilst our group showed that baseline plasma p-tau181 associates with cognitive decline and neurodegeneration one year later [15], it is unknown if baseline and serial measures of plasma p-tau181 can predict future progression to dementia in a larger cohort of individuals followed over longer periods. The relevance of plasma p-tau181 for monitoring the clinical and pathological progression of AD is unclear owing to the lack of longitudinal data. Addressing these knowledge gaps is critical for determining the suitability of using plasma p-tau181 for population screening, diagnosis, and as a recruitment and outcome measure for clinical trials [22].

In this study, we have investigated plasma p-tau181 in the Alzheimer’s Disease Neuroimaging Initiative (ADNI) cohort. We examined: (i) how plasma p-tau181 performs diagnostically in a large multicenter study to verify findings from recent single-center studies of plasma p-tau181 [15, 19, 20], (ii) how the biomarker performs in a head-to-head comparison with CSF biomarkers (p-tau181 and Aβ42), MRI and plasma NfL, (iii) if baseline plasma p-tau181 concentration is predictive of cognitive decline and conversion to AD dementia, and (iv) how longitudinal trajectories of plasma p-tau181 reflect stages of the AD *continuum*.

## Materials and methods

### Study participants

We used data from the multicenter ADNI study designed to develop and validate neuroimaging and biochemical biomarkers for the early detection, monitoring and treatment of AD [23]. This North American cohort recruited participants across 57 sites in the US and Canada. ADNI was launched in 2003, with clinical assessments and biospecimen collection from 7^th^ September, 2005 to 16^th^ June, 2016. The primary goal of ADNI has been to test whether serial MRI, PET, other biological markers, and clinical and neuropsychological assessments can be combined to measure the progression of mild cognitive impairment (MCI) and early AD. AD and MCI classification followed the criteria described elsewhere [23, 24]. The ADNI inclusion/exclusion criteria are described in detail at www.adni-info.org. Informed consent was provided by the enrolled participants or their authorized representatives. The ADNI study was approved by the local Institutional Review Boards at the participating centers. In addition, the present study was performed in accordance with the Strengthening the Reporting of Observational Studies in Epidemiology (STROBE) reporting guideline [25].

This study was based on participants with available plasma p-tau181 data (accessed on 20th June, 2020). The time at first plasma p-tau181 measurement defined the baseline time point of our study, which was used for the cross-sectional analyses, as well as the time of diagnostic classification. Longitudinal plasma p-tau181 data (for up to 96 months from baseline) was also evaluated. The number of time points varied between subjects, with the average being 3.11 (median= 3) visits per subject. For a detailed description of longitudinal data, see the Supplementary Appendix. Additional biomarkers assessing cognition, amyloid and tau pathologies as well as neurodegeneration were also investigated cross-sectionally and longitudinally even though they were available for subsets of the cohort, as described below.

### Plasma measurements

Blood samples were collected, processed, stored and analyzed as described previously [26, 27]. Plasma p-tau181 was measured using a clinically validated *in-house* assay described previously [15]. Plasma p-tau181 was measured on Simoa HD-X instruments (Quanterix, Billerica, MA, USA) in April 2020 at the Clinical Neurochemistry Laboratory, University of Gothenburg, Mölndal, Sweden, by scientists blinded to participants’ clinical information. Plasma p-tau181 data was collected over 47 analytical runs. Assay precision was assessed by measuring two different quality control samples at the start and end of each run, resulting in within-run and between-run coefficients of variation of 3.3%-11.6% and 6.4%-12.7%, respectively (Supplementary Table S1). A third quality control sample was used as internal calibrator. Out of 3762 ADNI plasma samples, four were not analyzed due to inadequate volumes. The remaining 3758 all measured above the assay’s lower limit of detection (0.25 pg/mL), with only six below the lower limit of quantification (1.0 pg/mL).

### Other biochemical measurements

CSF Aβ_42_, p-tau181 and t-tau were measured using the fully automated Elecsys assays (Roche Diagnostics) [28]. In this study, CSF data were matched with plasma biomarker data collected on the same study visit. Plasma NfL was measured in the same subjects as p-tau181 at the Clinical Neurochemistry Laboratory, University of Gothenburg, Mölndal, Sweden, using an *in-house* Simoa immunoassay, as previously described [26, 27].

### Cognition tests

Cognitive performance was assessed using the sum of boxes of the Clinical Dementia Rating (CDR-SOB), the Mini-Mental State Examination (MMSE) and the Alzheimer’s Disease Assessment Scale-Cognitive Subscale (ADAS-Cog). Cognitive scores were matched with plasma p-tau181 data based on the study visit.

### Neuroimaging

MRI and PET acquisitions followed ADNI protocols (http://adni.loni.usc.edu/methods). The MRI T1-weighted images underwent initial preprocessing with intensity normalization and gradient unwarping. They were processed using DARTEL and registered using a six-parameter affine transformation and nonlinearly spatially normalized to the ADNI template. PET images were pre-processed to have an effective point spread function of full-width at half-maximum of 8 mm. Subsequently, linear registration and nonlinear normalization to the ADNI template were performed with the transformations deriving from the automatic PET to MRI transformation and the individual’s anatomic MRI co-registration. Brain Aβ load was estimated using [^18^F]florbetapir (Aβ PET), tau load using [^18^F]flortaucipir (tau PET) and glucose uptake using [^18^F]fluorodeoxyglucose (FDG PET) standardized uptake value ratios (SUVR). SUVR volumes were generated using the full cerebellum as reference region for [^18^F]florbetapir, the cerebellar gray for [^18^F]flortaucipir, and cerebellar vermis and the pons as the reference regions for [^18^F]fluorodeoxyglucose. Summary PET measures include average SUVR of the meta-ROI regions for tau PET [29], the average SUVR of precuneus, cingulate, inferior parietal, medial prefrontal, lateral temporal, and orbitofrontal cortices for amyloid and the average of the bilateral angular gyrus, bilateral posterior cingulate, and bilateral inferior temporal gyrus for FDG PET [30]. Specific for tau PET, there was a large difference in time between first blood collection and the scan acquisition, and to account for this variability, we used the residuals of tau PET SUVR regressed on the time difference between the two measurements in the analysis.

Brain atrophy was estimated using hippocampal, whole brain and ventricular volumes. These measurements were estimated using FreeSurfer [31] and were adjusted for total intracranial volume (ICV), as previously described [32]. ICV adjustment was performed using data from all cognitively unimpaired (CU) subjects at baseline.

Here, we used the imaging data with the closest acquisition date to the plasma collection. Details on the number of subjects per PET modality at each time point can be found in the Supplementary Appendix.

### Cut-points

The study participants were further classified by their clinical diagnosis and Aβ status (Aβ+/-) defined by Aβ PET. The cut-off for Aβ PET (>1.11 SUVR) was determined using receiver operating characteristic (ROC) curve analysis contrasting AD versus CU (Youden index).The cut-off definition for CSF p-tau181 (>27 ng/L) has been previously described [26].

### Statistical analysis

All statistical analyses were performed in R statistical platform v.3.6.3 [33]. Demographic comparisons were done using chi-square test for categorical variables and one-way analysis of variance (ANOVA) followed by Tukey’s post-hoc test for continuous variables. Linear regression models (LM) tested the associations between plasma p-tau181 concentrations and other variables at baseline, always adjusting for age and gender. Cross-sectional group differences were also evaluated using linear regressions, adjusting again for age and sex, and Tukey’s honest significance test was used for post hoc pairwise comparisons, when necessary.

Longitudinal data were also evaluated with linear mixed-effect (LME) models, which always included random intercept and slopes and were adjusted for age, gender and baseline measures when needed. The models were fit using maximum likelihood estimation and time was set as continuous variable. We first compared plasma p-tau181 progression between categorical groups. Then, baseline plasma p-tau181 (continuous) was associated with longitudinal data for the other measures. The predictive power of baseline plasma p-tau181 (categorical) was evaluated by examining the difference between p-tau181 positive and negative groups in relation to cognitive decline and progressive neurodegeneration.

Biomarker rate of change was calculated using individual random slopes from LME models including random intercept and slopes and adjusted for age, gender. The rates of change were then correlated using Pearson’s correlation coefficient.

Receiver operating characteristic (ROC) curve analysis was used to assess diagnostic accuracies (CU vs AD) and to define the cut-off for plasma p-tau181. ROC 95% confidence intervals (CI) of sensitivities and specificities were also computed (Youden index).

Cox proportional hazard regression models tested the association between binarized values of plasma p-tau181 and the risk of incident AD dementia. The model was adjusted for age and sex and participants were censored at diagnosis of AD dementia or at their last follow-up visit. Hazard ratios (HR) were reported. Schoenfield residuals tested the assumption of proportional hazards and Martingale residuals assessed nonlinearity

The coefficients of variance (CV), and respective confidence intervals, were used to compare within-person p-tau181 variation over time. This analysis was performed using the individual average rate of change, which was calculated by subtracting baseline from follow-up (last visit available) plasma p-tau181 and dividing it by the time difference between the two points.

To facilitate comparison and interpretation of findings, LM and LME were performed using standardized variables when indicated. The Z-scores were based on the mean and standard deviation of the control population. Plasma p-tau181 and NfL were log transformed before standardization.

Were considered outliers subjects in which baseline plasma p-tau181 values were 3 standard deviations (SD) above or below the average of the whole population. These subjects (n=8) were excluded from the analyses.

## Results

### Demographic characteristics

In total, 1177 participants were included in the study, 1022 of whom having serial plasma collections resulting in a total of 3758 individual measures of plasma p-tau181. At baseline, 400 participants were clinically classified as CU, 558 as MCI and 219 with AD dementia. The mean age of the population was 74.1 years (SD=7.6), with MCI participants being younger on average than CU and AD (detailed demographic characteristics are presented in Supplementary Table 2). As expected, AD patients had worse performance in cognitive scores, increased load of Aβ and tau pathologies (evidenced by both CSF and imaging biomarkers), reduced brain metabolism (indexed by FDG PET) and increased brain atrophy as compared to CU and MCI participants. A total of 414 (41.4%) were Aβ+. When, stratifying by clinical syndrome, 68 (20%) of the CU, 209 (43%) of the MCI and 137 (77%) of the AD dementia individuals were Aβ+. Overall characteristics of this subsample are further described in Table 1.

**Table 1.** Baseline and longitudinal participant characteristics according to clinical diagnosis and Aβ PET status.

| Characteristic<sup>a,b</sup> | Aβ- CU<br>(n=268) | Aβ+ CU<br>(n=68) | Aβ- MCI<br>(n=277) | Aβ+ MCI<br>(n=209) | Aβ- AD<br>dementia<br>(n=41) | Aβ+ AD<br>(n=137) |
| --- | --- | --- | --- | --- | --- | --- |
| Age at baseline, years | 73.5(6.5) | 76.9(6.2) | 71.4(8.0)<sup>*,#</sup> | 73.9(6.7) | 77.3(7.0)<sup>*,#</sup> | 73.4(8.2) |
| Sex (female), No. (%) | 131(48.9%) | 43(63.2%) | 125(45.1%) | 87(41.6%) | 9(22.0%) | 66(47.8%) |
| $\geq 1$ APOE ε4, No. (%) | 61(22.8%)<sup>*</sup> | 33(48.5%)<sup>*,#</sup> | 90(32.3%)<sup>*</sup> | 142(67.9%)<sup>#</sup> | 14(34.1%)<sup>*</sup> | 108(78.3%)<sup>#</sup> |
| Educational level, years | 16.7(2.7)<sup>*</sup> | 16.6(2.2) | 16.3(2.6) | 15.9(2.8)<sup>#</sup> | 15.8(2.4) | 15.7(2.7)<sup>#</sup> |
| **CSF biomarkers at baseline, mean(SD)** |  |  |  |  |  |  |
| Aβ<sub>42</sub>, pg/mL | 1143.4(332.5)<sup>*</sup> | 744.4(253.8)<sup>*,#</sup> | 1078.9(353.9)<sup>*</sup> | 681.9(196.6)<sup>*,#</sup> | 867.8(392.9)<sup>*,#</sup> | 590.7(187.1)<sup>#</sup> |
| P-tau181, pg/mL | 18.0(7.4)<sup>*</sup> | 31.1 (12.4)<sup>*,#</sup> | 18.9 (8.7)<sup>*</sup> | 34.1 (13.8)<sup>*,#</sup> | 33.9(19.8)<sup>#</sup> | 39.9(15.5)<sup>#</sup> |
| Total-tau, pg/mL | 200.8 (73.8)<sup>*</sup> | 310.1 (115.2)<sup>*,#</sup> | 207.5 (86.6)<sup>*</sup> | 339.0 (125.8)<sup>*,#</sup> | 348.0 (180.7)<sup>#</sup> | 399.2 (149.6)<sup>#</sup> |
| **Imaging measure at baseline, mean(SD)** |  |  |  |  |  |  |
| Aβ PET | 0.9(0.1)<sup>*</sup> | 1.3(0.1)<sup>*,#</sup> | 1.0(0.1)<sup>*</sup> | 1.3(0.1)<sup>#</sup> | 1.0(0.1)<sup>*</sup> | 1.3(0.1)<sup>#</sup> |
| Hippocampal volume, mm<sup>3</sup> | 7609.3 (746.6)<sup>*</sup> | 7117.6 (763.1)<sup>*,#</sup> | 7114.1 (1077.6)<sup>*,#</sup> | 6655.9 (1025.3)<sup>*,#</sup> | 5709.0 (855.3)<sup>#</sup> | 5914.5 (887.6)<sup>#</sup> |
| Whole brain, mm<sup>3</sup> | 1053916.5 (61303.2)<sup>*</sup> | 1028401.0 (64418.9)<sup>*</sup> | 1055008.6 (66773.0)<sup>*</sup> | 1039169.6 (62083.3)<sup>*</sup> | 988329.7 (63365.8)<sup>#</sup> | 998562.3 (61268.6)<sup>#</sup> |
| Ventricular volume, mm<sup>3</sup> | 36000.8(17561.8)<sup>*</sup> | 39765.7 (17164.8)<sup>*</sup> | 39421.3 (20891.3)<sup>*</sup> | 39369.8 (18233.9)<sup>*</sup> | 57963.6 (25203.5)<sup>#</sup> | 47973.4 (18256.9)<sup>#</sup> |
| FDG-PET | 1.3(0.1)<sup>*</sup> | 1.3(0.1)<sup>*</sup> | 1.3(0.1)<sup>*</sup> | 1.2(0.1)<sup>*,#</sup> | 1.1(0.2)<sup>*,#</sup> | 1.0(0.1)<sup>#</sup> |
| **Cognitive score at baseline** |  |  |  |  |  |  |
| MMSE | 29.1(1.3)<sup>*</sup> | 28.8(1.0)<sup>*</sup> | 28.4(1.7)<sup>*,#</sup> | 27.6(1.9)<sup>*,#</sup> | 23.7(1.9)<sup>#</sup> | 22.9(2.7)<sup>#</sup> |
| CDR-SOB | 0.04(0.1)<sup>*</sup> | 0.2(0.4)<sup>*</sup> | 1.4(0.9)<sup>*,#</sup> | 1.7(1.1)<sup>*,#</sup> | 4.2(1.7)<sup>*,#</sup> | 4.8(2.0)<sup>#</sup> |
| ADAS-Cog | 5.3(3.0)<sup>*</sup> | 6.2(2.6)<sup>*</sup> | 8.0(3.8)<sup>*,#</sup> | 10.6(4.5)<sup>*,#</sup> | 19.8(7.0)<sup>*,#</sup> | 21.2(7.9)<sup>#</sup> |
| **Plasma biomarkers at baseline, mean(SD)** |  |  |  |  |  |  |
| Plasma p-tau181, pg/mL | 14.2(9.0)<sup>*</sup> | 19.1(8.2)<sup>*,#</sup> | 14.6(9.9)<sup>*</sup> | 22.8(9.9)<sup>#</sup> | 19.4(6.5)<sup>*</sup> | 25.5(8.6)<sup>#</sup> |
| Plasma NfL, pg/mL | 35.9(16.6)<sup>*</sup> | 42.4(16.1) | 38.2(23.9)<sup>*</sup> | 44.6(23.5)<sup>#</sup> | 50.9(21.0)<sup>#</sup> | 46.9(19.6)<sup>#</sup> |

<sup>a</sup> Continuous variables are given as mean (SD)<sup>b</sup> Aβ status defined by Aβ PET.<sup>\*</sup> p<0.05 compared with the Aβ + AD group<sup>#</sup> p<0.05 compared with the Aβ - CU group
Continuous variables were compared with one-way ANOVA followed by Tukey's post-hoc test. Categorical variables were compared with Chi test

Longitudinal plasma p-tau181 concentrations were available for 1022 individuals, including 361 CU (16% Aβ+), 508 MCI (38% Aβ+) and 153 AD dementia (67% Aβ+; Supplementary Table 3). The number of visits per participant ranged from 1 to 5, being the median 3 (CU=3; MCI=4; AD=2). As the majority of the available plasma p-tau181 data ranges between 0-48 months only these values were considered for longitudinal analysis (Supplementary Fig. 1). Further details on the number of participants with longitudinal biomarkers and cognitive measures are described in described in Supplementary Table 3.

### Plasma p-tau181 is increased along the Alzheimer’s *continuum*

In accordance with previous reports, baseline plasma p-tau181 concentrations were significantly higher in AD dementia (mean=23.6 pg/mL, SD=±8.8) and MCI (mean=18.3 pg/mL, SD=±10.8) as compared to the CU group (mean=14.9 pg/mL, SD=±9.0; *P*<0.0001; Fig. 1A), irrespective of Aβ status. The higher plasma p-tau181 concentration in AD dementia as compared to the MCI group was highly significant (*P*<0.0001).

**Fig. 1.**
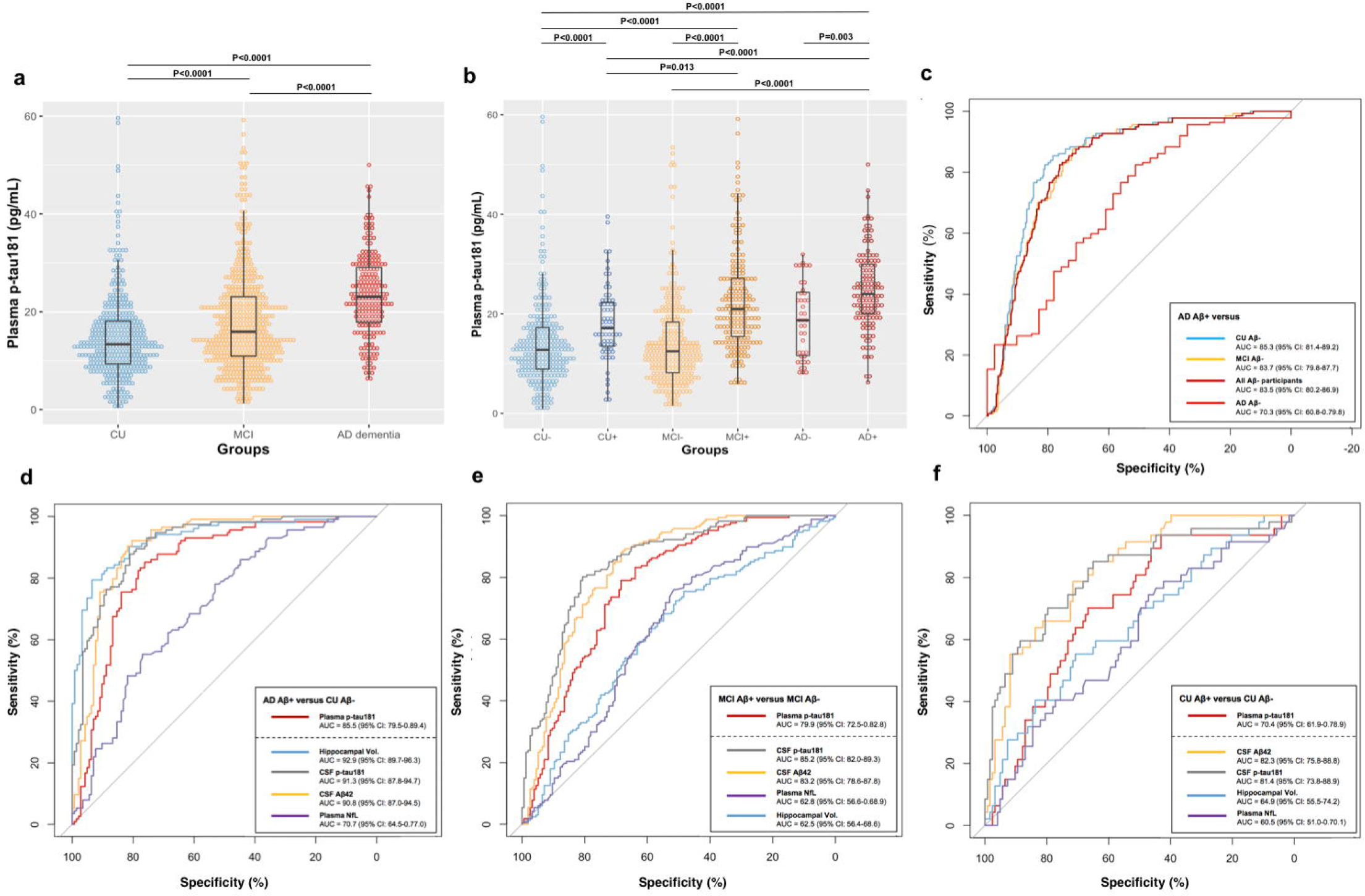
Plasma p-tau181 profile. Distribution of plasma p-tau181 concentrations across clinically defined diagnostics groups (a) showing higher biomarker levels associated with symptomatic disease stages. When considering Aβ PET status in addition to clinical diagnosis (b), plasma p-tau181 was found at higher concentrations in Aβ+ groups. The accuracy of plasma p-tau181 in distinguishing Aβ+ AD from other diagnostic groups is evidenced by AUCs as shown in (c). In addition, the accuracy of plasma p-tau181 in identifying Aβ pathology was evaluated in the context of other biomarkers (d-f).

When considering Aβ status (Fig. 1b), plasma p-tau181 was higher in Aβ+ CU, MCI Aβ+ and AD Aβ+ than Aβ-CU (all *P*<0.0001). Within the same clinical classification, plasma p-tau181 was higher in participants classified as Aβ+ as compared to those determined as Aβ- (CU, *P*<0.0001; MCI, *P*<0.0001; AD, *P*=0.003). Plasma p-tau181 was also increased in MCI Aβ+ and AD Aβ+ compared with Aβ+ CU (both *P*<0.0001).

### The diagnostic performance of plasma p-tau181

To test plasma p-tau181 accuracy in distinguishing clinically and biomarker-defined diagnostic groups, ROC tests compared Aβ+ AD dementia patients against all other groups (Fig. 1c). Plasma p-tau181 differentiated Aβ+ AD from Aβ- CU (AUC=85.3%; 95% CI, 81.4%-89.2%) and Aβ- MCI (AUC=83.8%; 95% CI, 79.8%-87.7%). Importantly, plasma p-tau181 distinguished Aβ+ MCI from Aβ- MCI (AUC=79.9%; 95% CI, 72.5%-82.8%) and also Aβ+ CU from Aβ- CU (AUC=70.4%; 95% CI, 61.9%-78.9%). In addition, plasma p-tau181 distinguished Aβ+ AD from Aβ- AD (AUC=70.3%; 95% CI, 60.8%-79.8%). Plasma p-tau181 separated Aβ+ AD from all Aβ- participants (AUC=83.5%; 95% CI, 80.2%-86.9%). Using a cut-off value of 17.7 pg/mL (generated from ROC analyses, comparing Aβ+ AD dementia and Aβ- CU) classified 44.1% (n=521) of the participants as positive for plasma p-tau181. Analysis of concordance showed that 28.5% (n=285) of the participants were positive concordant with Aβ PET and 27.4% (n=245) concordant with CSF p-tau181 (Supplementary Fig. 2). Participants who tested negative for plasma p-tau181 were 55.8% (n=659) of the study population. Concordant negative cases were 42.8% (n=428) and 41.4% (n=367) in relation to Aβ PET and CSF p-tau181, respectively. Discordant data were 28.7% (n=286) in relation to Aβ PET and 31.2% (n=279) in relation to CSF p-tau181. A cut point to define Aβ positivity based on plasma p-tau181 was also calculated by comparing individuals positive and negative for Aβ PET. Applying a cut-off value of 14.5 pg/mL (AUC=76.9%; 95% CI, 74.0%-79.8%) classified 42.1% of the CU, 56.1% of MCI and 85.4% of the AD participants as positive for Aβ pathology.

Next, we evaluated the accuracy of plasma p-tau181 to identify Aβ+ at differing stages of the Alzheimer’s *continuum* and compared them to reference standard plasma, CSF and imaging biomarkers, in the same participants. Firstly, and as stated above, plasma p-tau181 had a good performance in separating Aβ+ AD dementia and Aβ+ CU with comparable AUCs as compared with CSF p-tau181, CSF Aβ_42_ and hippocampal volume (Fig. 1d). Plasma p-tau181 outperformed plasma NfL in this comparison (Fig. 1d). We then tested the performance of plasma p-tau181 to identify Aβ+ in CU and MCI groups. In this analysis, plasma p-tau181 demonstrated a significantly higher AUC than plasma NfL in identifying Aβ+ participants at both stages (Fig. 1e-f), and also in identifying misclassified AD *e.g*., Aβ- AD (p-tau181, AUC=70.3; NfL, AUC=54.9). At the CU, MCI and dementia stages, plasma p-tau181 also demonstrated higher AUCs in identifying Aβ pathology than hippocampal volume. Plasma p-tau181 had marginally lower AUCs in identifying Aβ+ cases compared with CSF p-tau181 and CSF Aβ_42_ at both the CU and MCI stages. At the dementia stage, plasma p-tau181 was superior to CSF p-tau181 (AUC=62.6; 95% CI, 48.9%-76.4%) and comparable to CSF Aβ_42_ (AUC=72.1; 95% CI, 59.7%-84.4%) in predicting Aβ pathology.

### Plasma p-tau181 associates cross-sectionally with CSF, plasma, imaging and cognitive biomarkers

At the cross-sectional level, plasma p-tau181 correlated with CSF p-tau181 (r=0.36, *P*<2.2×10^−16^), Aβ_42_ (=-0.39, *P*<2.2×10^−16^) and t-tau (r=0.33; *P*<2.2×10^−16^), as well as with plasma NfL (r=0.39; *P*<2.2×10^−16^; Supplementary Fig. 3a-h). Increased concentrations of plasma p-tau181 were also correlated with high Aβ (r=0.42; *P*<2.2×10^−16^) and tau PET SUVR (r=0.26; *P*<2.9×10^−5^) and reduced FDG PET SUVR (r=-0.3; *P*<2.2×10^−16^). Brain atrophy was also correlated with plasma p-tau181 as indexed by hippocampal volume (r=-0.34; *P*<2.2×10^−16^), ventricular volume (r=0.24; *P*<2.2×10^−16^) and total brain volume (r=-0.23; *P*<2.2×10^−16^). Further, worse performance in cognitive assessments was correlated with higher plasma p-tau181 concentrations (r_MMSE_=-0.3; r_ADAS-Cog_=0.34; r_CDR-SOB_=0.31; all *P*<2.2×10^−16^).

Linear models were applied to evaluate the association between plasma p-tau181 and the above-mentioned biomarkers, but now adjusting for age, sex and diagnosis (Fig. 2a-h). After adjusting for covariates, higher plasma p-tau181 levels were associated with higher CSF p-tau181 (*t*=8.78, *P*=2×10^−16^) and t-tau concentrations (*t*=7.59, *P*= 7.64×10^−14^) and lower CSF Aβ_42_ levels (*t*=-8.27, *P*= 6.11×10^−16^) as expected. In addition, high plasma p-tau181 was associated with high brain Aβ and tau load (*t* _Aβ PET_=10.39, *P* _Aβ PET_ =2×10^−16^; *t* _tau PET_=4.07, *P* _tau PET_ =6.15×10^−5^) whilst inversely associated with FDG PET (*t*=-4.91, *P*=1.02×10^−6^). Plasma p-tau181 was associated with measures of brain atrophy (hippocampal volume, *t*=-4.47, *P*=8.48×10^−6^; ventricular volume, *t*=3.18, *P*=1.10×10^−3^), except for total brain volume that was no longer associated with plasma p-tau181 after adjusting for covariates (*t*=-1.72, *P*=0.08). Worse cognition was also associated with higher levels of plasma p-tau181 (*t* _MMSE_=-4.864, *P* _MMSE_ =1.34×10^−6^; *t*_ADAS-Cog_=5.936, *P*_ADAS-Cog_ =4.03×10^−9^; *t*_CDR-SOB_=5.604, *P* _CDR-SOB_ = 2.71×10^−8^). All linear models described here were also applied within diagnostic groups and results are summarized in Supplementary Table S4.

**Fig. 2.**
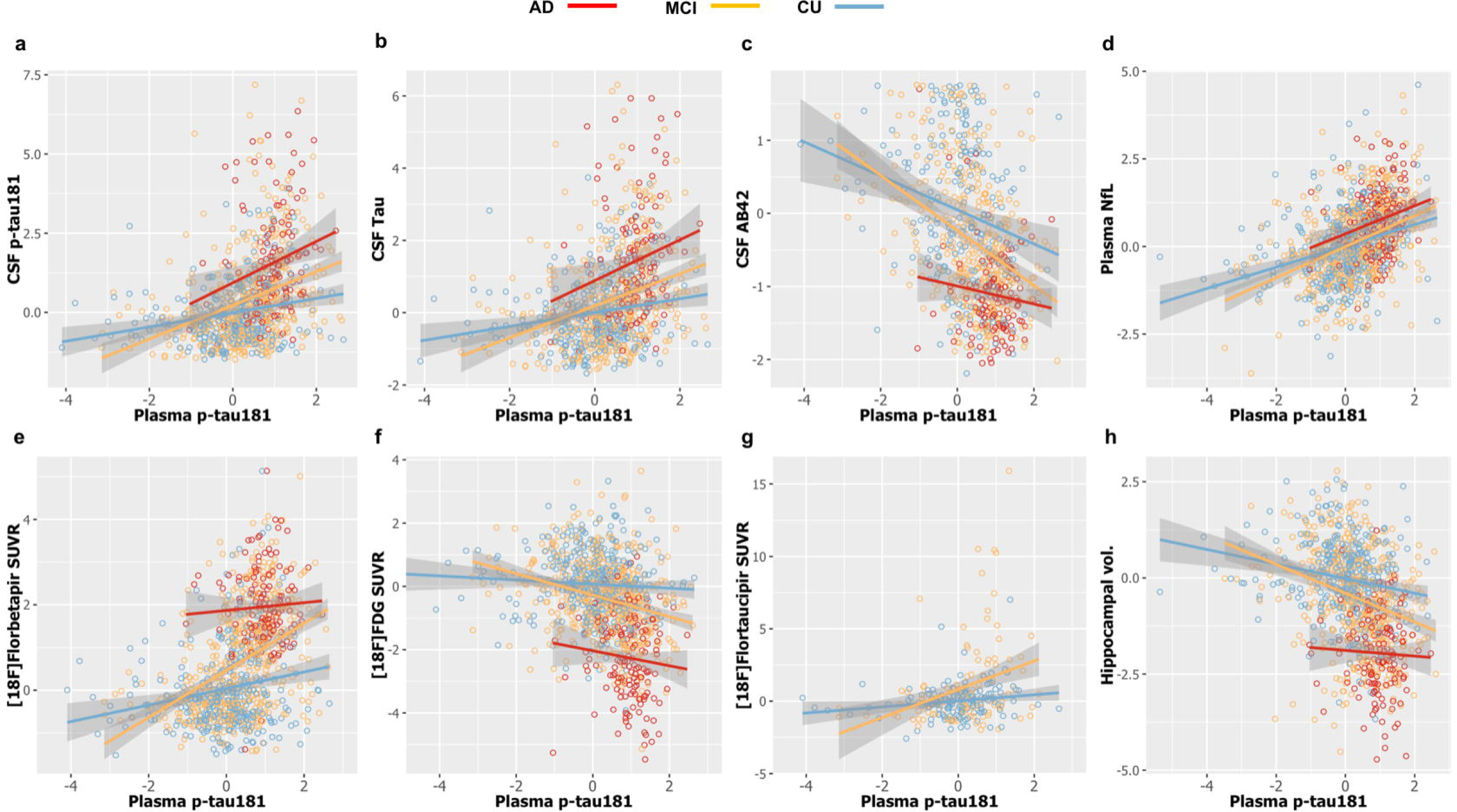
Cross-sectional associations at baseline. Linear regression analyses show that plasma p-tau181 is significantly associated with CSF, plasma and imaging biomarkers within diagnostic groups (a-h). Detailed description of findings are reported in Supplementary Table S4.

Linear models also compared the association between plasma p-tau181 and all other biomarkers between groups defined according to Aβ PET positivity (Supplementary Fig. 4a-h). Overall, plasma p-tau181 was more strongly associated with worsened phenotypic presentations in the Aβ+ compared with the Aβ- group (detailed results in Supplementary Table S5).

### Baseline plasma p-tau181 predicts progression to dementia, faster cognitive decline and worsening neurodegeneration

Survival analysis evaluated the risk of progression to AD dementia considering one’s baseline plasma p-tau181 status (plasma p-tau181 concentrations >17.7 pg/mL were considered positive) and clinical diagnosis. The analysis included 729 participants (283 CU and 446 MCI) with baseline plasma and CSF p-tau181 and up to 84 months of diagnosis data. High plasma p-tau181 was associated with increased risk of AD dementia in MCI (hazard ratio [HR]=22.75, 95% CI, 9.90-52.31; Fig. 3b) and CU Aβ+ (HR=3.25, 95% CI, 1.12-9.40) as compared with Aβ- CU. This was seen to be similar to the associations found using CSF p-tau181 (HR _Aβ+MCI_=37.1, 95% CI, 15.0-91.8; HR _Aβ+CU_=5.4, 95% CI, 1.8-16.3, Fig. 3a).

**Figure 3.**
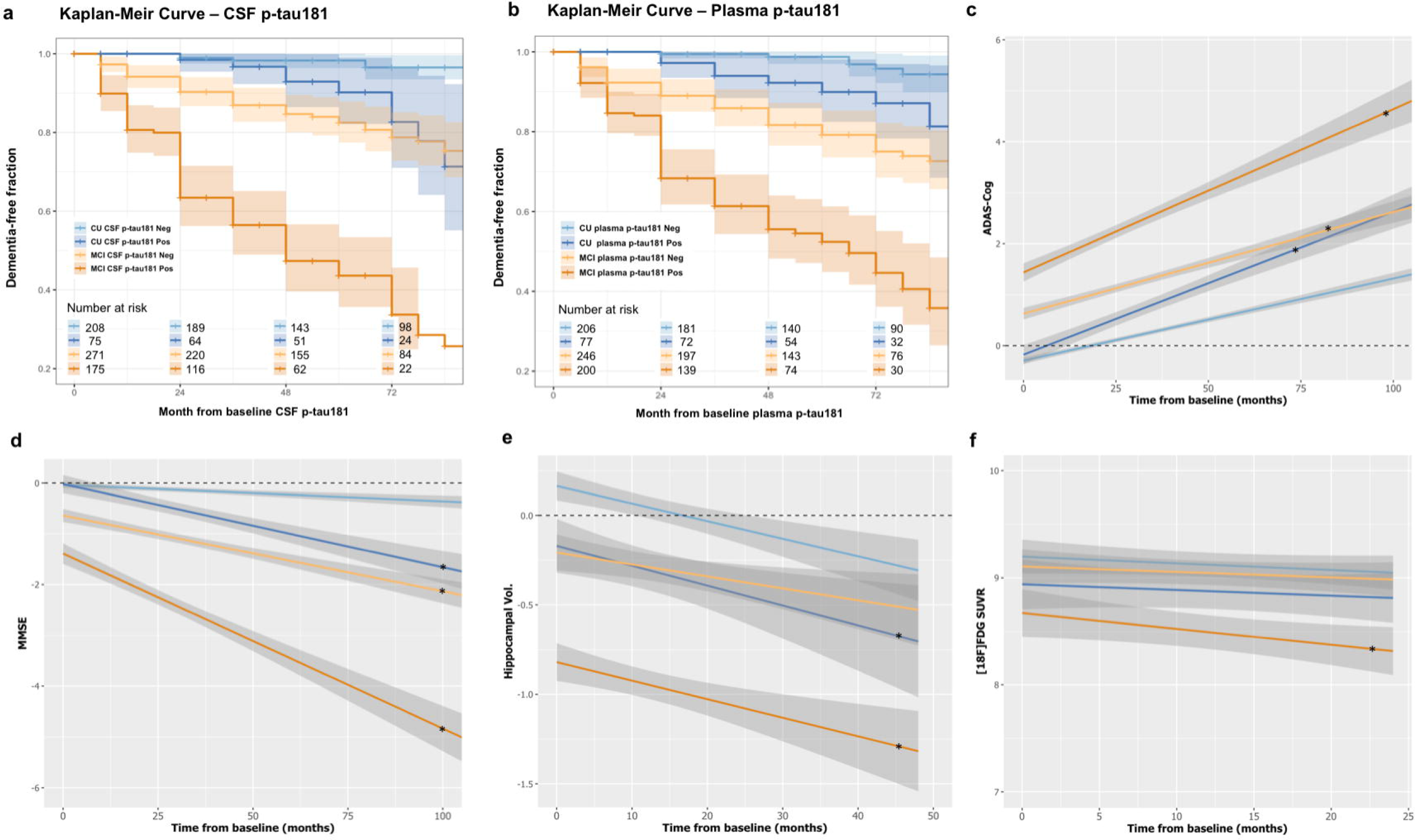
Plasma p-tau181 as a predictor. Cox-proportional hazard model showing that higher levels of baseline CSF (a) and plasma (b) p-tau181 are associated with an increased risk to progress to AD dementia, as evidenced by the Kaplan-Meier curves. Moreover, linear mixed effect models indicated that high baseline plasma p-tau181 levels are associated with worse cognitive performance (c-d) and faster neurodegenerative processes (e-f). The * indicates the longitudinal trajectories that are significantly different from plasma p-tau181 negative CU (the reference group).

When evaluating the predictive power of plasma p-tau181 to detect changes in downstream biomarkers of pathological progression in CU and MCI, LME analysis showed that participants who were plasma p-tau181-positive at baseline, and free from dementia, had faster cognitive decline over 100 months in comparison to CU p-tau181-negative individuals in two cognitive tests evaluated here (Fig. 3c-d; for detailed description of the results see Supplementary Table S6). Similarly, higher rates of hippocampal atrophy (over 48 months) were observed in MCI plasma p-tau181-positive at baseline (*t*=-6.07; *P*=2.15×10^−9^) and CU plasma p-tau181-positive at baseline (*t*=-2.15; *P*=0.03), but not MCI plasma p-tau181-negative (*t*=-1.14; *P*=0.25), compared with those who were CU plasma p-tau181-negative (Fig. 3e). In addition, faster decline in FDG PET uptake over 24 months was observed in plasma p-tau181-positive MCI participants (*t*=-6.07; *P*=2.15×10^−9^) when compared with CU plasma p-tau181-negative (Fig. 3f). Interestingly, when adding AD dementia subjects to the analysis, one can observe that MCI subjects who were positive for plasma p-tau181 reached follow-up biomarker levels consistent with baseline biomarker levels of the AD dementia group (Supplementary Fig. 5).

### Longitudinal characteristics of plasma p-tau181

Longitudinal plasma p-tau181 was first analyzed by computing individual slopes (up to 48 months), with the LME model accounting for age and sex. No significant difference was observed between average slopes of diagnostic groups (*t*=-0.34, *P*=0.92). Similarly, LME compared plasma p-tau181 progression over 24 months between diagnostic groups (also further classified according to Aβ PET status), and no significant difference was found between the slopes of the groups (*t*_CU+/CU-_=0.46; *t*_MCI-/CU-_=0.58; *t*_MCI+/CU-_=-0.42; *t*_AD+/CU-_=0.89; all *P*>0.05; Fig. 4a).

**Fig. 4.**
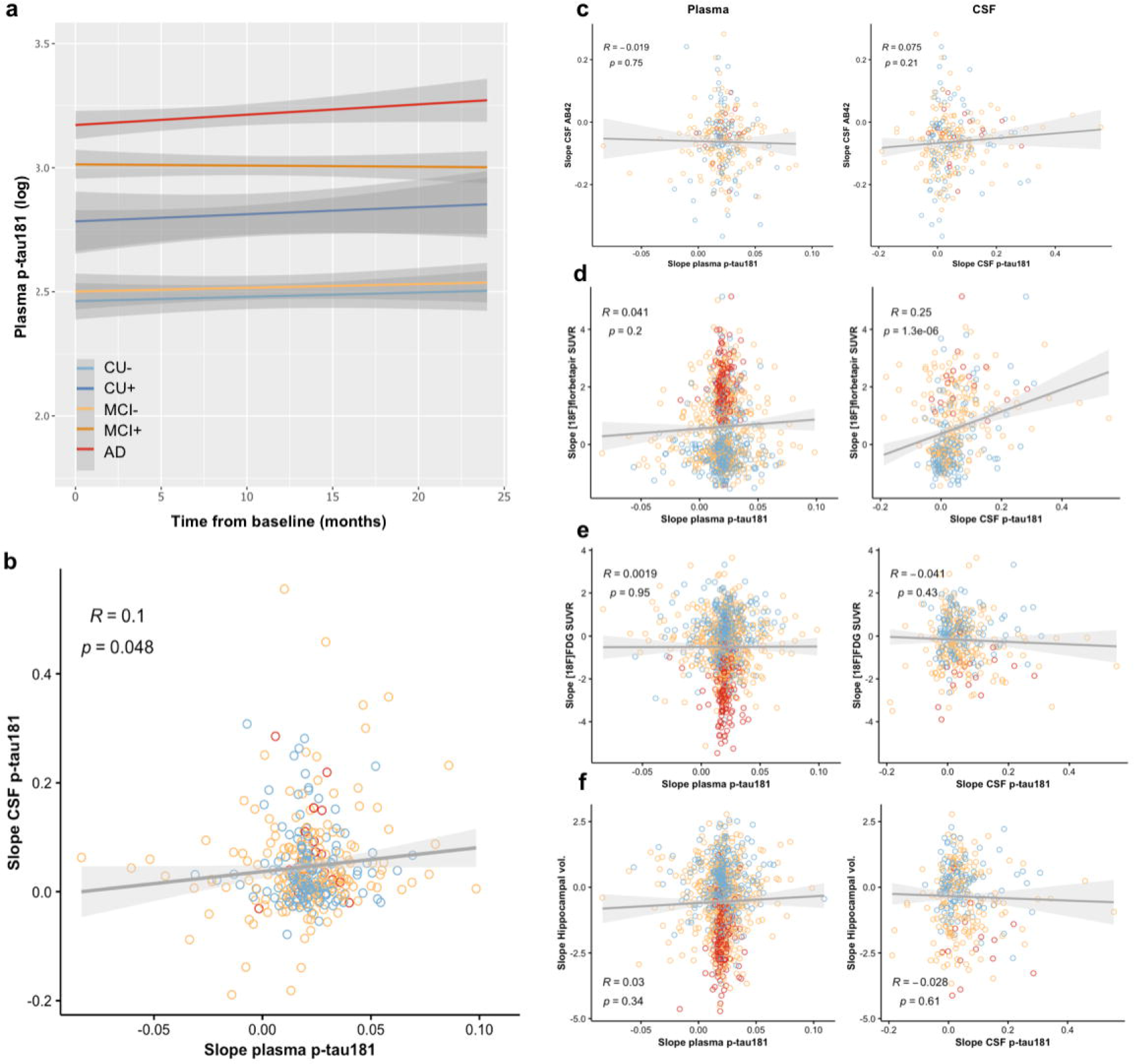
Longitudinal plasma p-tau181 profile. Linear mixed effect models indicate that plasma p-tau181 trajectories over time are similar across diagnostic groups (a). Pearson correlation analysis shows that individual plasma p-tau181 changes over time and are correlated with CSF p-tau181 changes (b). In addition, correlation plots (c-f) suggest comparable associations between CSF and plasma p-tau181 and the other CSF and imaging biomarkers.

To further evaluate the longitudinal relationship between plasma p-tau181 and relevant biomarkers, individual slopes were computed for all variables (adjusting for age and sex), and these values were then correlated with plasma p-tau181 slopes. A positive correlation was found between the slope of CSF p-tau181 and the slope of plasma p-tau181 (*r*=0.1, *P*=0.04; Fig 4b). No significant correlation was found between longitudinal changes in plasma p-tau181 and the other biomarkers evaluated (Fig 4c-f). Importantly, when comparing longitudinal changes in CSF p-tau181 with changes in the other biomarkers, similarly to plasma p-tau181, no significant correlations were found, with the exception for Aβ PET (Fig 4c-f; r=0.25, *P*=1.3×10^−6^).

To evaluate biomarker stability over time, the average rate of change (Supplementary Fig. 6) was calculated with raw plasma p-tau181 values and this was then used to calculate the coefficient of variance (CV) for each of the diagnostic groups. Plasma p-tau181 was highly stable over the study period, with very low within-person variability (CV: CU=7%, MCI=9%, AD=12%). The CVs were slightly higher in the respective Aβ+ cases in the MCI and AD dementia groups (Aβ- CU (7%, range=6.4%-7.6%); Aβ+ CU (6%, range=5.2%-7.5%); Aβ- MCI (7%, range=6.5%-7.5%); Aβ+ MCI (12%, range=10.9%-13.3%); Aβ- AD dementia (10%, range=7.8%-13.9%) and Aβ+ AD (14%, range=12.3%-16.3%)). Similar results were found when only considering changes over 24 months, which would reflect the typical time window for therapy in clinical trials: Aβ- CU (9%, range=8.4%-10.1%), Aβ+ CU (7%, range=5.9%-8.6%), Aβ- MCI (9%, range=8.2%-9.8%), Aβ+ MCI (12%, range=10.9%-13.3%), Aβ- AD (10%, range=7.8%-13.9%) and Aβ+ AD (14%, range=12.3%-16.3%).

## Discussion

In this multicenter study, performed in 1177 participants from the ADNI cohort, we verify and confirm the findings from recent single-center cohort studies [15, 19, 20] that plasma p-tau181 (1) is higher in AD dementia and is increased along the Alzheimer’s *continuum*, (2) identifies Aβ+ irrespective of disease stage, (3) correlates with CSF p-tau181 and (4) predicts future progression to AD dementia, cognitive decline and hippocampal atrophy. Moreover, utilising the longitudinal aspect of ADNI, we have demonstrated that serial sampling of p-tau181 remains moderately stable overtime, with low individual variability; the small changes detected overtime are significantly related to longitudinal changes in CSF p-tau181 but not any other biomarker modalities.

The significance of the results from this multicentre study, and those from single centre studies that have preceded it, have potentially two major implications. Firstly, the clinical identification of AD dementia could be greatly aided by plasma p-tau181 in primary care, which is important given that discrepancies are common between the clinical diagnosis of AD [34]. In the National Institute of Aging and Alzheimer Association (NIA-AA) Research Framework [35], AD is now defined as a biological construct, documented by biomarker evidence of AD pathology (that is, evidence of both Aβ and tau pathology), and not as a clinical syndrome; thus, plasma p-tau181 could be implemented as a cost-effective and rapid tool to triage possible cases of AD in primary care that would be referred to specialised centers. Moreover, plasma p-tau181 could be used in population-based studies to detect individuals at high risk to develop AD and enrolled them in clinical trials. Plasma p-tau181 has shown to have very high accuracy in determining AD from non-AD neurodegenerative diseases (*e.g*., frontotemporal dementia, progressive supranuclear palsy and cortico-basal syndromes) which is comparable to tau PET [15, 19–21]. Although this particular comparison could not be performed in this AD-focused study due to low numbers of subjects with matching plasma p-tau181 and tau PET results, we have shown that individuals who have been clinically defined as AD dementia or MCI but lack biomarker evidence of Aβ pathology have significantly lower plasma p-tau181 concentration. Plasma p-tau181 could also identify Aβ pathology in these cognitively impaired groups. This shows great promise that in this clinically challenging scenario, where clinical symptoms are seemingly identical and CSF/PET biomarkers are not available, plasma p-tau181 would provide valuable information to a clinician, which could improve the confidence in an AD diagnosis and administering symptomatic treatment (*e.g*., acetylcholinesterase inhibitors or memantine), better inform on patient management, and in the future guide in the selection of patients eligible for disease-modifying treatments, if such reach the clinic. Moreover, if plasma p-tau181 does not indicate AD, despite cognitive decline, then further investigation could be warranted and implemented at a far earlier stage *e.g*., FDG PET, dopamine transporter (DAT) or for MRI for frontotemporal dementia, dementia with Lewy bodies or vascular dementia, respectively [36]. This could be accompanied by plasma NfL as increases in this biomarker would indicate on-going neurodegeneration, irrespective of the underlying pathology [26, 27, 37].

An additional clinical application of plasma p-tau181 is the prediction of progression to dementia in individuals with MCI or CU Aβ+. We have demonstrated that plasma p-tau181 accurately predicts the development of dementia, which had a similar predictive performance as CSF p-tau181. This verifies previous findings from the Swedish BioFINDER study [19] but in a multicentric fashion. We also dichotomized participants by MCI and CU to show that individuals with a positive plasma p-tau181 at baseline are still more likely to develop dementia even if no cognitive impairment is present. Importantly, CU individuals with a negative value of plasma p-tau181 at baseline show almost no progression to dementia over a 7-year period. These findings are further supported by high plasma p-tau181 at baseline being associated with deterioration in cognitive function as assessed with neuropsychiatric batteries routinely used in primary care and specialist facilities. We further showed that Aβ+ individuals (MCI or CU) deteriorated at significantly faster rate than the respective Aβ- individuals. Together, these results support that baseline plasma p-tau181 is highly predictive of future AD diagnosis and AD-mediated cognitive decline.

The second implication would be the use of plasma p-tau181 for participant selection or monitoring in trials that are targeting AD-related brain pathologies or in large-scale epidemiological and genetic studies to identify novel risk and resilience factors for AD. As previously discussed, we and others have shown the ability to demonstrated the presence of Aβ pathology at the MCI stage with relatively good accuracy (AUC=79.9%), which was only slightly inferior to CSF Aβ_42_ and CSF p-tau181 (AUC=83.2%, AUC=85.2%, respectively) but vastly outperformed plasma NfL and MRI measures of hippocampal volume (AUC<66%). This has fundamental importance in the design of therapeutic trials targeting individuals at the symptomatic phase of the disease, where a positive plasma p- tau181 test could confirm AD or reduce screening failure rate even if individuals fulfil the clinical criteria for AD dementia or MCI.

The accuracy of plasma p-tau181 to identify Aβ+ subjects at the asymptomatic stage was lower, despite highly significant differences from Aβ- participants, and it was evident that CSF Aβ_42_ and CSF p-tau181 were significantly more sensitive to identify brain Aβ pathology. While this points towards plasma p-tau181 having greater utility at the symptomatic stage of the disease than to detect preclinical pathology, other studies show that plasma p-tau181 robustly increase in the preclinical phase in familial AD mutation carriers [38]. Nonetheless, plasma p-tau181 could still act as a pre-screening aid in clinical trials to enrich an asymptomatic population for greater success by a secondary investigation (*e.g*., Aβ PET). The large multicentric design of this study allowed for speculation on the cost-benefit analysis of plasma p-tau181 as pre-screening for clinical trials targeting the unimpaired population. Of the 336 CU participants included in this study, 68 demonstrated Aβ-positivity, as indexed by [^18^F]florbetapir, resulting in a positivity rate of 20% in the asymptomatic sub-cohort – 10% lower than previously reported prevalence estimates [39, 40]. When employing our cut-off for Aβ positivity (14.5 pg/mL), we classify 143 CU participants as “*p-tau181-positive for A*β” of which 60/68 were actually Aβ+ by PET. Considering the prevalence shown in this study, a typical trial design aiming to recruit 1000 Aβ+ asymptomatic individuals would require ∼5000 individuals scanned at the cost of $15,000,000 (assuming a cost of 3000 $/scan). On the contrary, a pre-screening design using p-tau181 in 5000 participants (estimated $50/participant) would cost $250,000 and would yield 2125 plasma p-tau181-positive tests. These 2125 individuals could subsequently be scanned by Aβ PET at an estimated cost of $6,645,000 to confirm positivity for trial enrolment. Comparatively, the pre-screening approach would save in the region of $8,300,000, which is over half of the original cost by employing Aβ PET alone. At this preclinical stage, the addition of plasma Aβ_42_/Aβ_40_ to p-tau181 could be an additional tool that improves the accuracy Aβ+ prediction and therefore reduce costs further [19]. In addition to being economically advantageous, plasma p-tau181 pre-screening would be time-saving and also logistically simpler to recruit 5000 individuals (or more) willing to undergo blood sampling compared with PET imaging.

We observed longitudinal stability of plasma p-tau181 over several years, which demonstrates that plasma p-tau181 has low biological variability, and measures are methodologically stable and reliable across repeated samplings. This observation could also be of potential benefit in disease-modifying trials seeking a measurable response to a therapeutic target. In fact, when CSF p-tau181 concentrations from individuals with paired CSF and plasma data were analysed, similar observations were made, including a lack of significant longitudinal association with other biomarkers, although a significant but weak correlation existed for CSF p-tau181 and [^18^F]florbetapir longitudinal change. These findings are supported by a previous report on the ADNI cohort that showed that it would take at least 6.2 years for CSF p-tau181 concentrations to significantly start altering [41]. Changes in plasma would potentially require longer duration since blood is downstream of CSF with respect to central nervous system metabolites. However, we did demonstrate that the longitudinal slopes of plasma and CSF p-tau181 were significantly correlated suggesting, to some degree, that the subtle changes in CSF are reflected in blood. Toledo and colleagues [41] also reported that CSF p-tau181 increased up to 5.1 pg/mL per year. As the plasma p-tau181 concentration is around 5% of that in CSF [15], an increase of around 0.26 pg/mL per year in blood would be seen. The longitudinal trajectories recorded in the present study were similar to these values.

This study is not without limitations. The study lacked sufficient tau PET data at the time of blood collection. Although CSF p-tau181 is an highly accurate diagnostic biomarker for AD, tau PET better reflects the degree of NFT pathology [42] and is a superior diagnostic tool for AD [36]. We also used Aβ PET to classify +/- groups. There was a relatively common discordance between CSF and PET biomarkers, meaning that individuals classified as Aβ- by PET could be Aβ+ by CSF. Previous reports have demonstrated that changes in Aβ CSF precede alterations in Aβ PET [43], and therefore individuals classified as Aβ- in this study could have underlying and developing Aβ pathology. Furthermore, high rates of dropout in the AD group precluded extensive analyses of the long-term trajectories of plasma p-tau181 in the most advanced stages of the disease (> 48 months). Additionally, the potential effect of comorbidities such as vascular dementia contributing on plasma p-tau181 could not be examined. However, vascular dementia is unlikely to confound plasma p-tau181 measures since patients with this form of dementia have low concentrations of both CSF and plasma p-tau181 [15].

In summary, plasma p-tau181 is a promising and accurate diagnostic and prognostic biomarker for AD, particularly when CSF or PET examination is not possible. We have also shown that plasma p-tau181 is encouraging for clinical trial use and can be utilised in symptomatic or asymptomatic populations to considerably lower costs to enrich a population prior to Aβ PET confirmation. Furthermore, the longitudinal, within-person plasma p-tau181 measures were shown to be stable over four years demonstrating a potential utility to evaluate and monitor the effects of novel disease-modifying treatments.

## Data Availability

The files used in preparing this manuscript are publicly available from http://adni.loni.usc.edu/. All data is available in the main text or the supplementary materials.

## Acknowledgments

Data collection and sharing was funded by ADNI (NIH #U01 AG024904) and DOD ADNI (#W81XWH-12-2-0012). ADNI is funded by the National Institute on Aging, the National Institute of Biomedical Imaging and Bioengineering, and through generous contributions from the following: AbbVie, Alzheimer’s Association; Alzheimer’s Drug Discovery Foundation; Araclon Biotech; BioClinica, Inc.; Biogen; Bristol-Myers Squibb Company; CereSpir, Inc.; Cogstate; Eisai Inc.; Elan Pharmaceuticals, Inc.; Eli Lilly and Company; EuroImmun; F. Hoffmann-La Roche Ltd and its affiliated company Genentech, Inc.; Fujirebio; GE Healthcare; IXICO Ltd.; Janssen Alzheimer Immunotherapy Research & Development, LLC.; Johnson & Johnson Pharmaceutical Research & Development LLC.; Lumosity; Lundbeck; Merck & Co., Inc.; Meso Scale Diagnostics, LLC.; NeuroRx Research; Neurotrack Technologies; Novartis Pharmaceuticals Corporation; Pfizer Inc.; Piramal Imaging; Servier; Takeda Pharmaceutical Company; and Transition Therapeutics. The Canadian Institutes of Health Research is providing funds to support ADNI clinical sites in Canada. Private sector contributions are facilitated by the Foundation for the National Institutes of Health (www.fnih.org). The grantee organization is the Northern California Institute for Research and Education, and the study is coordinated by the Alzheimer’s Therapeutic Research Institute at the University of Southern California. ADNI data are disseminated by the Laboratory for Neuro Imaging at the University of Southern California.

TKK holds a Brightfocus fellowship (#A2020812F), and is further supported by the Swedish Alzheimer Foundation (Alzheimerfonden; #AF-930627), the Swedish Brain Foundation (Hjärnfonden; #FO2020-0240), the Swedish Dementia Foundation (Demensförbundet), the Agneta Prytz-Folkes & Gösta Folkes Foundation (#2020-00124), the Aina (Ann) Wallströms and Mary-Ann Sjöbloms Foundation, the Anna Lisa and Brother Björnsson’s Foundation, Gamla Tjänarinnor, and the Gun and Bertil Stohnes Foundation. NJA is supported by the Swedish Alzheimer Foundation (Alzheimerfonden; #AF-931009), the Swedish Brain Foundation (Hjärnfonden), the Agneta Prytz-Folkes & Gösta Folkes Foundation, and the Swedish Dementia Foundation (Demensförbundet). AS was supported by the Emil Aaltonen Foundation and the Paulo Foundation, and currently receives funding from the Orion Research Foundation. MS-C received funding from the European Union’s Horizon 2020 Research and Innovation Program under the Marie Sklodowska-Curie action grant agreement No 752310, and currently receives funding from Instituto de Salud Carlos III (PI19/00155) and from the Spanish Ministry of Science, Innovation and Universities (Juan de la Cierva Programme grant IJC2018-037478-I). PR-N is supported by the Weston Brain Institute, the Canadian Institutes of Health Research, the Canadian Consortium on Neurodegeneration in Aging and the Fonds de Recherche du Québec – Santé (FRQS; Chercheur Boursier, and 2020-VICO-279314 TRIAD/BIOVIE Cohort), the CIHR-CCNA Canadian Consortium of Neurodegeneration in Aging, and the Canada Foundation for Innovation (project 34874). KB was supported by the Alzheimer Drug Discovery Foundation (ADDF; #RDAPB-201809-2016615), the Swedish Research Council (#2017-00915), the Swedish Alzheimer Foundation (#AF-742881), Hjärnfonden, Sweden (#FO2017-0243), and a grant (#ALFGBG-715986) from the Swedish state under the agreement between the Swedish government and the County Councils, the ALF-agreement. KB is supported by the Swedish Research Council (#2017-00915), the Alzheimer Drug Discovery Foundation (ADDF), USA (#RDAPB-201809-2016615), the Swedish Alzheimer Foundation (#AF-742881), Hjärnfonden, Sweden (#FO2017-0243), the Swedish state under the agreement between the Swedish government and the County Councils, the ALF-agreement (#ALFGBG-715986), and European Union Joint Program for Neurodegenerative Disorders (JPND2019-466-236). HZ is a Wallenberg Scholar supported by grants from the Swedish Research Council (#2018-02532), the European Research Council (#681712), Swedish State Support for Clinical Research (#ALFGBG-720931), the Alzheimer Drug Discovery Foundation (ADDF), USA (#201809-2016862), the European Union’s Horizon 2020 research and innovation programme under the Marie Skłodowska-Curie grant agreement No 860197 (MIRIADE), and the UK Dementia Research Institute at UCL.

## Author contributions

TKK, ALB, NJA, PSC, PR-N, KB and HZ conceptualized the research; TKK, NJA, JLR, AS, MS-C, HK, UA, KB and HZ performed plasma p-tau181 measurements, data quality control and data compilation; TKK, ALB, NJA, FL, PSC, AMR, MS, TAP, PR-N, KB, and HZ contributed to data analysis; ALB, FL, PSC, TAP and PR-N developed and implemented algorithms for data analysis; TKK, ALB, NJA, KB, and HZ wrote the original manuscript draft. All authors reviewed, edited and approved the final manuscript for submission.

